# Striatal Dopamine Binding in Anhedonia: A Simultaneous [^11^C]Raclopride Positron Emission Tomography and Functional Magnetic Resonance Imaging Investigation

**DOI:** 10.1101/2022.07.21.22277878

**Authors:** Rachel D. Phillips, Erin C. Walsh, Nicole R. Zürcher, David Lalush, Jessica Kinard, Chieh-En Tseng, Paul Cernasov, Delia Kan, Kaitlin Cummings, Lisalynn Kelley, David Campbell, Daniel G. Dillon, Diego A. Pizzagalli, David Izquierdo-Garcia, Jacob Hooker, Moria Smoski, Gabriel S. Dichter

## Abstract

**Background:** Anhedonia is hypothesized to be associated with blunted mesocorticolimbic dopamine (DA) functioning in samples with major depressive disorder. The purpose of this study was to examine linkages between striatal DA binding, reward circuitry functioning, anhedonia, and, in an exploratory fashion, self-reported stress, in a transdiagnostic anhedonic sample.

**Methods:** Participants with (n=25) and without (n=12) clinically impairing anhedonia completed a reward-processing task during simultaneous positron emission tomography and magnetic resonance (PET-MR) imaging with [^11^C]raclopride, a DA D2/D3 receptor antagonist that selectively binds to striatal DA receptors.

**Results:** Relative to controls, the anhedonia group exhibited increased [^11^C]raclopride binding potential (ΔBP_ND_) in response to rewards, interpreted as decreased task-related DA release, in the left putamen, caudate, and nucleus accumbens and right putamen and pallidum. There were no group differences in task-related brain activation (fMRI) during reward processing after correcting for multiple comparisons. General functional connectivity (GFC) findings revealed blunted fMRI connectivity between PET-derived striatal seeds and target regions (i.e., bilateral caudate, putamen, pallidum, medial prefrontal cortex, anterior cingulate cortex, and thalamus) in the anhedonia group. Associations were identified between anhedonia severity and the magnitude of task-related DA release to rewards in the left putamen, but not mesocorticolimbic GFC. We did not find evidence of associations between self-reported stress and striatal DA response to rewards, mesocorticolimbic fMRI activation or GFC in the anhedonic sample.

**Conclusions:** Results provide evidence for reduced striatal DA functioning during reward processing and blunted mesocorticolimbic network functional connectivity in a transdiagnostic sample with clinically significant anhedonia.

## Introduction

Anhedonia is characterized by impaired reward processing and blunted mesocorticolimbic dopamine (DA) system functioning (1–3). This ascending DA tract passes through reward learning (-meso), cognitive control (-cortico), and emotional (-limbic) hubs of the brain (4), and impairments in motivation and the anticipation of rewards are associated with alterations in striatal DA tone, DA release, and DA signaling (3,5–7). Associations between anhedonia and mesocorticolimbic DA system functioning have primarily been investigated in major depressive disorder (MDD) (8,9). While anhedonia is a core symptom of MDD, it is also a transdiagnostic symptom that is pervasive across numerous neuropsychiatric disorders (10). A putative neural mechanism of anhedonia is striatal hypoactivation, and anhedonia severity negatively correlates with ventral striatal activity during the anticipation of rewards in depressed populations (11–13). Anhedonia severity is also associated with altered intrinsic functional connectivity between striatal regions and areas of the prefrontal cortex (PFC) in adolescents (12) and adults (14,15). In a non-clinical adult sample, reduced nucleus accumbens response to reward was uniquely related to anhedonia severity, and not depressive or anxious symptoms (16). Together these findings demonstrate distinct patterns of mesocorticolimbic DA system activation and connectivity associated with anhedonia.

Simultaneous positron emission tomography and magnetic resonance (PET-MR) imaging using [^11^C]raclopride, a radioligand that allows for the quantification of DA D2/D3 receptor binding, has demonstrated that functional magnetic resonance imaging (fMRI) activation and functional connectivity in mesolimbic brain regions during reward anticipation correlate with ventral striatal DA release in MDD (17) and non-clinical (18) samples. Anhedonia is associated with altered DA functioning, including decreased striatal DA transporter availability in MDD (9) and increased striatal DA D2/D3 receptor availability in MDD (8), although no association between anhedonia and DA release capacity in MDD has been reported (19). This inconsistency may be explained, in part, by the diagnostic heterogeneity of MDD as opposed to sampling an anhedonic phenotype.

Additionally, alterations in DA signaling, transmission, and reward circuitry functioning are associated with stress. Stress is believed to induce anhedonia via downregulation of mesocorticolimbic DA system functioning (3,7,20,21). In rodents, following chronic stress, DA release in the nucleus accumbens is inhibited, and this inhibition is associated with learned helplessness, anhedonic behaviors, and coping failure (22). In particular, uncontrollable and unpredictable stressors lead to the development of anhedonic-like phenotypes in animals (23– 25). Stress is also associated with impaired reward processing characterized by reduced goal-directed behavior, motivation, and reward responsiveness in clinical samples (3,26,27). Behaviorally, self-reported stress is also associated with blunted incentive motivation and altered reward anticipation (3,28). However, no research has examined associations between self-reported stress, anhedonia, and striatal dopamine functioning in a transdiagnostic anhedonic sample.

In the present study, we used simultaneous PET-MR imaging with the D2/D3 dopamine receptor antagonist [^11^C]raclopride in a transdiagnostic sample of adults with clinically impairing anhedonia. Our goal was to investigate relationships between anhedonia, striatal DA release, and mesocorticolimbic network functioning during reward processing. We hypothesized that the transdiagnostic anhedonia group would be characterized by decreased striatal task-related DA release to rewards, indexed by the non-displaceable binding potential (ΔBP_ND_) of [^11^C]raclopride, relative to a control group. We also hypothesized that striatal DA functioning would predict anhedonia severity. Next, we predicted that the anhedonia group would show decreased mesocorticolimbic network activation and connectivity during reward processing using fMRI. Finally, an exploratory aim was to examine associations between self-reported stress, anhedonia, and mesocorticolimbic DA system functioning. This aim was exploratory given that participants were not recruited based on stress exposure. We hypothesized that greater self-reported stress would be inversely associated with striatal DA release and mesocorticolimbic network fMRI activation and connectivity during reward processing.

## Methods

### Study Overview

The present study complements an ongoing NIMH-funded clinical trial (R61/R33 MH110027) investigating the effects of a novel psychosocial anhedonia treatment on neural responses to rewards and anhedonia symptoms (ClinicalTrials.gov Identifiers NCT02874534 and NCT04036136). Data from control participants, recruited as part of a separate study, have been reported previously (29). These companion studies met research standards for Institutional Review Board (IRB) approval at UNC-Chapel Hill and Duke University, and PET imaging protocols were approved by the UNC Radioactive Drug Research Committee. PET-MR imaging data acquisition occurred within four weeks of completing inclusion and exclusion assessment for the companion study. Written informed consent was obtained prior to inclusion in the study.

### Participants

#### Eligibility Criteria

Eligible participants in the ANH group were 18 to 50 years old, treatment-seeking for clinically significant anhedonia (i.e., Snaith-Hamilton Pleasure Scale (SHAPS) scores greater than or equal to 20 using the ordinal scoring of Franken and colleagues (30) and with Clinician’s Global Impression Scale Severity (CGI-S; (31) scores greater than or equal to 3, indicating clinical impairment). Eligible participants in the CON group had no present or past psychiatric diagnoses, as assessed by the Structured Clinical Interview for DSM-5 (SCID-5-RV) (32). Additional eligibility criteria are provided in Supplemental Materials IA.

Twenty-eight ANH participants and 23 CON participants completed PET-MR scans. Three ANH participants and 11 CON participants were excluded due to problems with the PET injection or scanner (4 CON participants), PET infusion (2 ANH participants), or technical errors at the scan (1 ANH and 7 CON participants). The final sample included 25 ANH participants and 12 CON participants.

### Clinical Diagnostic & Symptom Measures

The SCID-5-RV was used to assess eligibility and for clinical characterization. As previously reported, the control group had no lifetime psychiatric diagnoses, as assessed by the SCID-5-RV (29). Only participants in the ANH group completed the following self-report measures assessing stress and anhedonia severity.

The Perceived Stress Scale (PSS-10) was the primary measure of self-reported stress. The PSS assesses self-reported unpredictable and uncontrollable stressors over the past month and contains 10 items (33). Total scores range from 0 to 40, whereby higher scores indicate greater perceived stress (34).

The posttraumatic stress disorder (PTSD) Checklist (PCL-5), the secondary measure of self-reported stress, was used to assess PTSD symptoms in the last month. The PCL-5 is a well-validated scale with 20 items (35). Total scores range from 0 to 80, whereby higher scores indicate greater severity of symptoms. The PCL-5 version used in the current study did not include the Criterion A component. Therefore, scores reflect general distress in relation to stressful life events rather than a Criterion A trauma (36).

The Snaith–Hamilton Pleasure Scale (SHAPS) was the primary measure of anhedonia. The SHAPS is a well-validated 14-item questionnaire that assesses hedonic capacity. Total scores range from 14 to 56, whereby higher scores indicate greater anhedonia severity in the present state (i.e., “the last few days”).

The 21-item Beck Depression Inventory (BDI-II) was administered to assess depression symptom severity. Total scores range from 0 to 63, whereby higher scores indicate greater depressive severity. The BDI-II Anhedonia Subscale was used as a secondary measure of anhedonia. This comprises four items from the BDI-II (i.e., loss of interest, loss of pleasure, loss of interest in sex, and loss of energy) (37). Whereas the SHAPS primarily assesses aspects of consummatory reward, or pleasure, the BDI-II anhedonia subscale captures aspects of both consummatory and anticipatory reward processing (37).

### Neuroimaging Data

#### Simultaneous PET-MR scan protocol

Participants completed a 75-minute simultaneous PET-MR scan on a Siemens Biograph mMR scanner. A bolus+infusion protocol (*Figure 1*) was implemented for PET-MR scanning, using the D2/D3 antagonist [^11^C]raclopride, which selectively binds to striatal DA receptors (38). Dynamic PET acquisition for [^11^C]raclopride used a planned K_bol_ of 105 min, administered using a Medrad® Spectris Solaris® EP MR Injection System. See *Figure 1* for timing of data collection by modality.

**Figure 1.**
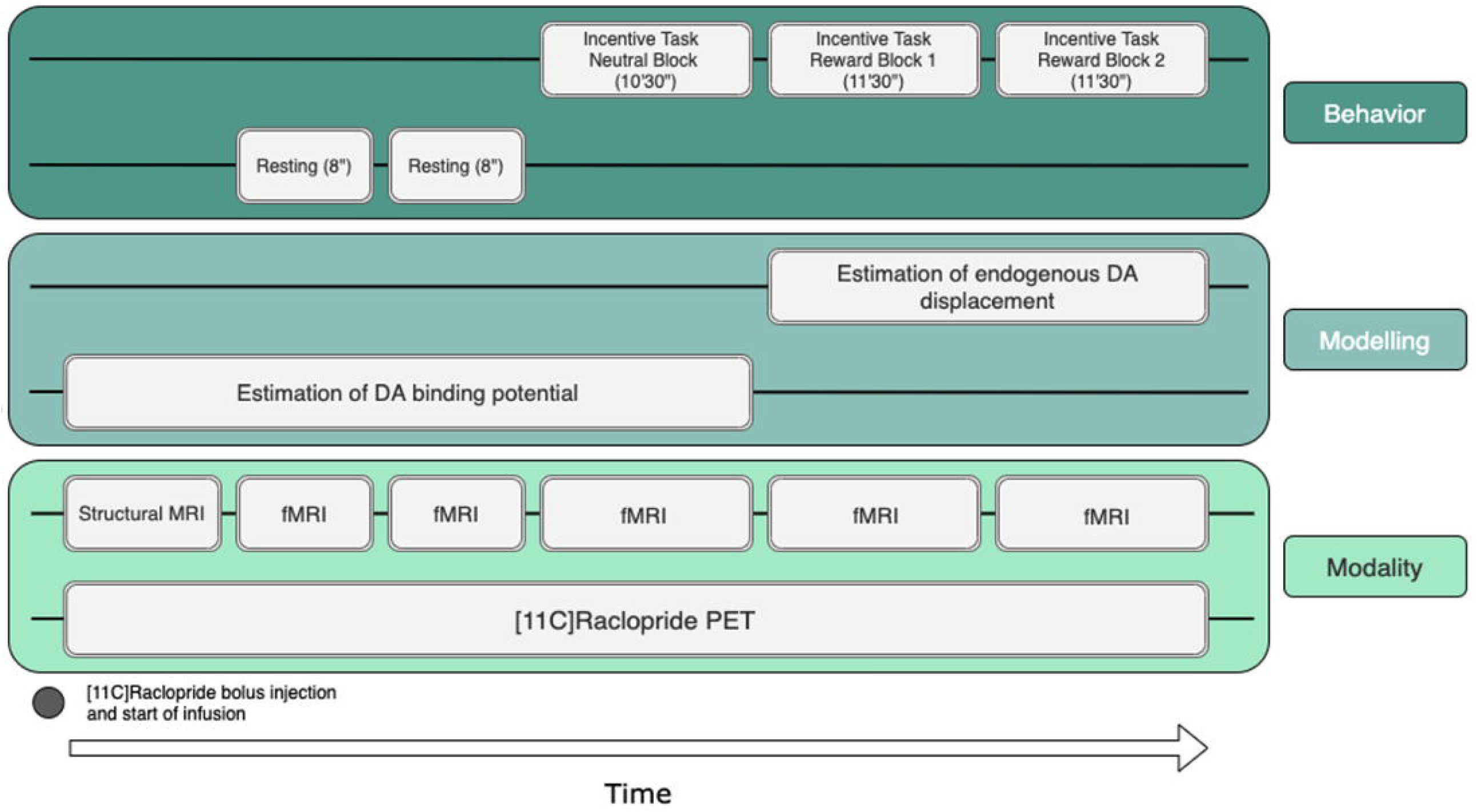
Timing of data collection, data modelling, and participant behavior during scanning. Three task blocks were presented during which fMRI data were collected simultaneously with the PET acquisition.

#### Reward task during PET and fMRI scanning

The reward task used during scanning is described in Supplemental Materials IC and illustrated in Figure 2.

**Figure 2.**
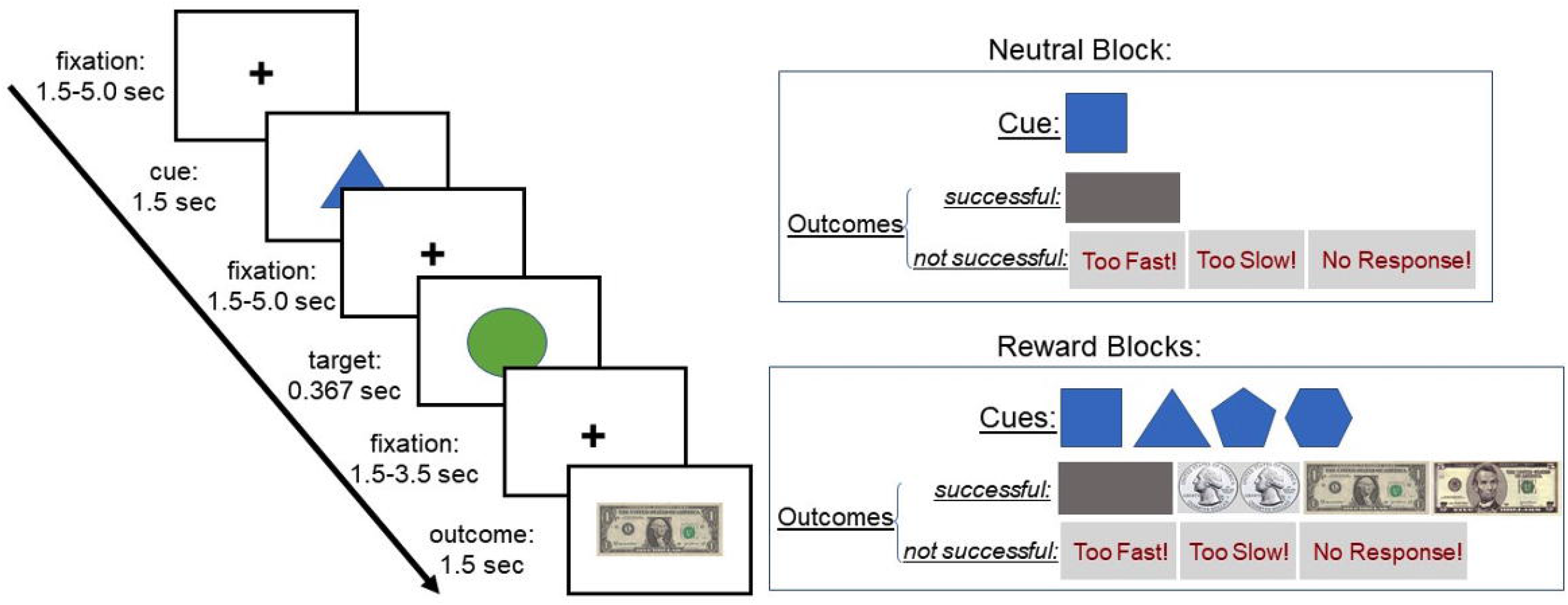
PET-MR Monetary Incentive Delay (MID) Task. Each trial consisted of a cue phase and an outcome phase. Trials were presented first in a neutral block that consisted of only neutral trials and then in two reward blocks that consisted of reward trials of varying magnitudes (small, medium, or large). The relationship between cue identity and outcome magnitude had to be learned by experience. Further details of the PET-MR Monetary Incentive Delay (MID) Task are provided in the Supplemental Materials IC.

#### PET Analysis

[^11^C]Raclopride is a D2/D3 receptor antagonist, and therefore competes with endogenous DA for receptors. Binding potential (BP_ND_), the ratio of selectively bound ligand to non-displaceable ligand in the tissue at equilibrium, was estimated from dynamic PET images for the neutral and reward blocks of the MID task per subject. BP_ND_ was quantified using the two-part simplified reference tissue model (SRTM) (39). Here, reward blocks encompass trials during which participants both anticipated and received rewards. Baseline BP_ND_ and change in BP_ND_ following reward task onset (Δ BP_ND_ %) measures DA functioning during the non-reward state and activation (reward task-related) states, respectively. This approach measures the extent to which endogenous DA displaces the radiotracer. A typical, or adaptive, DA response to rewards in the striatum would be indicated by lower BP_ND_ values during reward, relative to neutral, indicating that DA has increased and out-competed the tracer for binding sites (8). Accordingly, decreased ΔBP_ND_ is interpreted as increased task-related DA release.

To identify regions that showed between-group differences in ΔBP_ND_ from neutral to reward phases of the MID, for each subject, we estimated striatal DA functioning during each condition of the task. Here, we would expect negative ΔBP_ND_ values (Reward – Neutral) for controls if DA competes the tracer. A *z*-score statistical map representing the difference between groups and conditions (ANH - CON; Reward -Neutral) was created from subject images by contrasting voxel-wise ΔBP_ND_ (Reward - Neutral) maps. Thus, z-scores compare ΔBP_ND_ for the ANH group to ΔBP_ND_ for the CON group in such a way that positive z-scores indicate that anhedonic participants demonstrate less response in the expected direction than control participants (i.e., ΔBP_ND_ for the ANH group is less negative than ΔBP_ND_ for the CON group). This *z*-score statistical map was then thresholded at *z* > 2.58 (uncorrected) and anatomically constrained to the striatum (i.e., bilateral caudate, putamen, pallidum, and nucleus accumbens) using masks from the Harvard-Oxford probabilistic atlas.

For each significant functionally-defined cluster that emerged from this contrast, condition-specific ΔBP_ND_ values were extracted per participant. To study the pattern of results in greater detail, these values were then compared by evaluating group (ANH, CON)□×□condition (reward, neutral) interactions via analyses of variance (ANOVAs). For a complete description of PET analyses see Supplemental Materials IB and (40).

#### fMRI Image Preprocessing & Motion

See Supplemental Materials ID for additional details about fMRI Image Preprocessing & Motion.

#### fMRI Activation Analysis

To examine fMRI responses during reward anticipation, BOLD responses to reward cues of all magnitudes (small, medium, and large) vs. neutral cues were examined from cue onset to the end of the fixation period (i.e., during the cue and the target). To examine fMRI responses during reward processing, activation to successful vs. unsuccessful outcomes on reward trials of all magnitudes (small, medium, and large) were examined. See Supplemental Materials IF for additional details about fMRI activation analyses.

#### fMRI Connectivity Analysis

A general functional connectivity (GFC) approach examined whole-brain connectivity using striatal PET-derived seed regions that displayed significant differences in ΔBP_ND_ for the contrast ANH - CON; Reward - Neutral of the MID task. GFC, a method that combines resting-state and task fMRI data, offers better test-retest reliability and higher estimates of heritability than intrinsic connectivity estimates from the same amount of resting-state data alone (41). In the current study, the combination of two resting-state runs and three MID task blocks yielded approximately 45 minutes of fMRI data for connectivity analyses. This is critical given that >25 min of fMRI data are needed to reliably detect individual differences in connectivity (42). Voxel-wise whole-brain connectivity was evaluated using the CONN Toolbox’s seed-to-voxel analysis. Analyses corrected for multiple comparisons using a false-discovery rate (FDR) approach, at the familywise error (FWE) rate of *p* < .05.

#### Associations between PET-MR, Anhedonia, and Self-Reported Stress in the Anhedonia Group

To examine whether anhedonia severity and self-reported stress were associated with striatal DA function and mesocorticolimbic network functioning within the ANH group, we conducted statistical regression models in R, version 4.0.3 (43). Because only participants in the ANH group completed self-report measures assessing stress and anhedonia severity, analyses were limited to this group. PET-derived striatal ΔBP_ND_ and network functional connectivity values (i.e., fMRI-derived correlations between network regions with correlated BOLD signal change) were tested as individual predictors of anhedonia severity (i.e., SHAPS and BDI anhedonia subscale), in separate regressions. Additional regressions tested whether self-reported stress (i.e., PSS and PCL-5) predicted the magnitudes of these PET- and fMRI-derived variables. Corrections for multiple comparisons were made within each set of hypotheses (i.e., correcting across regression analyses that examined whether striatal DA release to rewards, or ΔBP_ND_, predicted anhedonia severity).

Lastly, bivariate Pearson correlations between clinical and PET-MR variables of interest were explored. Corrections for multiple comparisons were made within each set of analyses (i.e., clinical variables with striatal ΔBP_ND_ values and clinical variables with network functional connectivity values) using the false-discovery rate (FDR) method (44).

## Results

### Participant Characteristics

*Table 1* summarizes demographic information and descriptive statistics for the samples. *Table 2* reports clinical characteristics for the ANH group; the self-report measures assessing stress and anhedonia severity were not collected in the CON group.

**Table 1.**
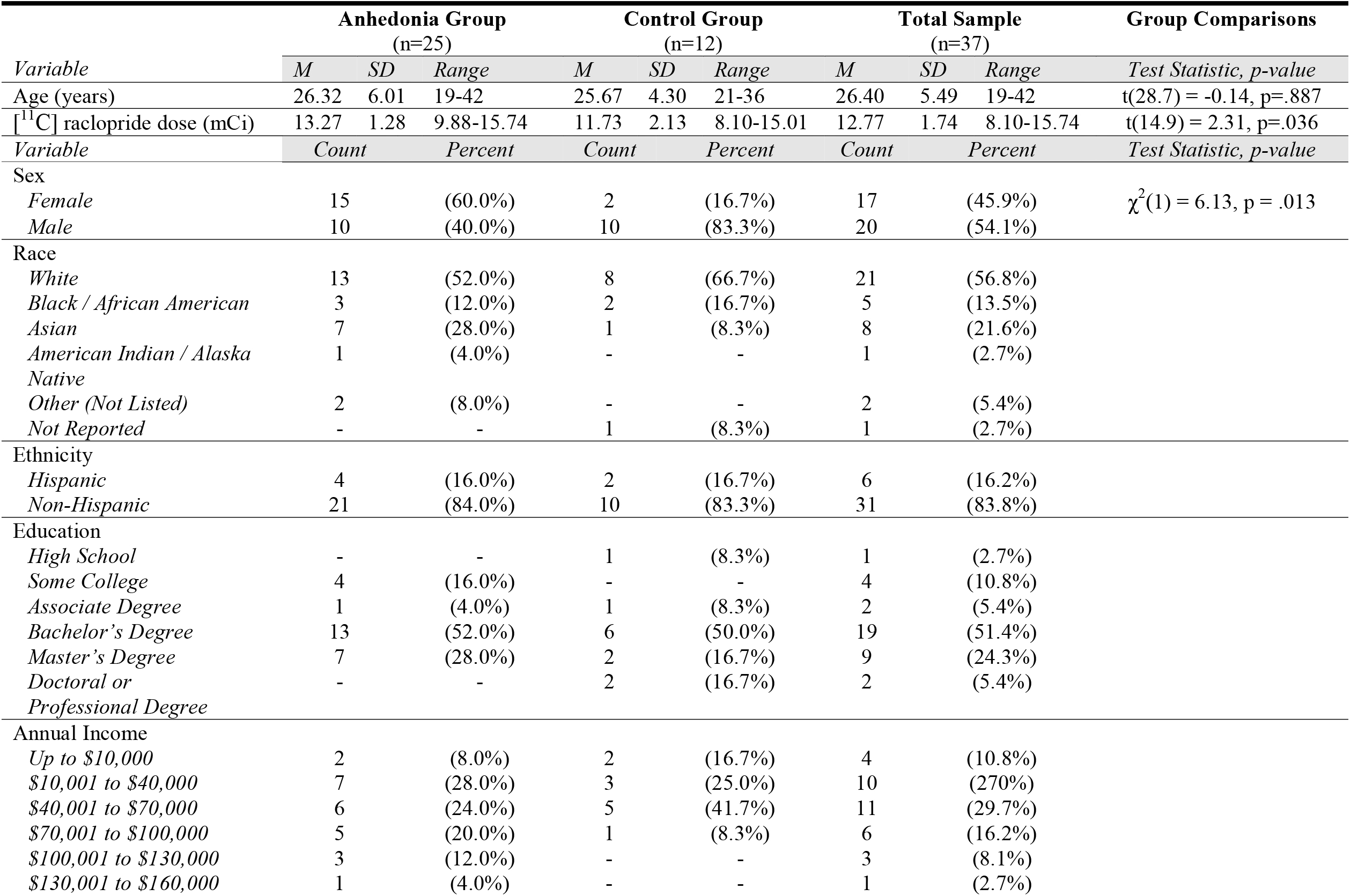
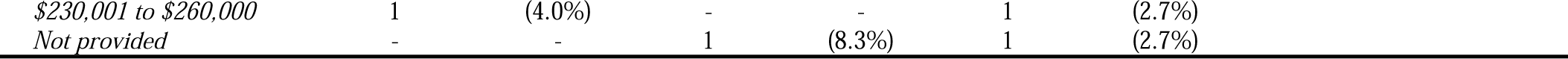
Sample Characteristics. Note – Participants were able to endorse one or more race categories.

|  | Anhedonia Group<br>(n=25) |  |  | Control Group<br>(n=12) |  |  | Total Sample<br>(n=37) |  |  | Group Comparisons |
| --- | --- | --- | --- | --- | --- | --- | --- | --- | --- | --- |
| Variable | M | SD | Range | M | SD | Range | M | SD | Range | Test Statistic, p-value |
| Age (years) | 26.32 | 6.01 | 19-42 | 25.67 | 4.30 | 21-36 | 26.40 | 5.49 | 19-42 | t(28.7) = -0.14, p=.887 |
| [<sup>11</sup>C] raclopride dose (mCi) | 13.27 | 1.28 | 9.88-15.74 | 11.73 | 2.13 | 8.10-15.01 | 12.77 | 1.74 | 8.10-15.74 | t(14.9) = 2.31, p=.036 |
| Variable | Count | Percent |  | Count | Percent |  | Count | Percent |  | Test Statistic, p-value |
| Sex |  |  |  |  |  |  |  |  |  |  |
| Female | 15 | (60.0%) | | 2 | (16.7%) | | 17 | (45.9%) | | $\chi^2(1) = 6.13, p = .013$ |
| Male | 10 | (40.0%) |  | 10 | (83.3%) |  | 20 | (54.1%) |  |  |
| Race |  |  |  |  |  |  |  |  |  |  |
| White | 13 | (52.0%) |  | 8 | (66.7%) |  | 21 | (56.8%) |  |  |
| Black / African American | 3 | (12.0%) |  | 2 | (16.7%) |  | 5 | (13.5%) |  |  |
| Asian | 7 | (28.0%) |  | 1 | (8.3%) |  | 8 | (21.6%) |  |  |
| American Indian / Alaska Native | 1 | (4.0%) |  | - | - |  | 1 | (2.7%) |  |  |
| Other (Not Listed) | 2 | (8.0%) |  | - | - |  | 2 | (5.4%) |  |  |
| Not Reported | - | - |  | 1 | (8.3%) |  | 1 | (2.7%) |  |  |
| Ethnicity |  |  |  |  |  |  |  |  |  |  |
| Hispanic | 4 | (16.0%) |  | 2 | (16.7%) |  | 6 | (16.2%) |  |  |
| Non-Hispanic | 21 | (84.0%) |  | 10 | (83.3%) |  | 31 | (83.8%) |  |  |
| Education |  |  |  |  |  |  |  |  |  |  |
| High School | - | - |  | 1 | (8.3%) |  | 1 | (2.7%) |  |  |
| Some College | 4 | (16.0%) |  | - | - |  | 4 | (10.8%) |  |  |
| Associate Degree | 1 | (4.0%) |  | 1 | (8.3%) |  | 2 | (5.4%) |  |  |
| Bachelor's Degree | 13 | (52.0%) |  | 6 | (50.0%) |  | 19 | (51.4%) |  |  |
| Master's Degree | 7 | (28.0%) |  | 2 | (16.7%) |  | 9 | (24.3%) |  |  |
| Doctoral or Professional Degree | - | - |  | 2 | (16.7%) |  | 2 | (5.4%) |  |  |
| Annual Income |  |  |  |  |  |  |  |  |  |  |
| Up to \$10,000 | 2 | (8.0%) | | 2 | (16.7%) | | 4 | (10.8%) | | |
| \$10,001 to \$40,000 | 7 | (28.0%) | | 3 | (25.0%) | | 10 | (27.0%) | | |
| \$40,001 to \$70,000 | 6 | (24.0%) | | 5 | (41.7%) | | 11 | (29.7%) | | |
| \$70,001 to \$100,000 | 5 | (20.0%) | | 1 | (8.3%) | | 6 | (16.2%) | | |
| \$100,001 to \$130,000 | 3 | (12.0%) | | - | - | | 3 | (8.1%) | | |
| \$130,001 to \$160,000 | 1 | (4.0%) | | - | - | | 1 | (2.7%) | | |

|  |  |  |  |  |  |  |
| --- | --- | --- | --- | --- | --- | --- |
| *\$230,001 to \$260,000* | 1 | (4.0%) | - | - | 1 | (2.7%) |
| *Not provided* | - | - | 1 | (8.3%) | 1 | (2.7%) |

**Table 2.** Anhedonia Group Clinical Characteristics. PSS – Perceived Stress Scale; SHAPS – Snaith-Hamilton Pleasure Scale; BDI-II – Beck Depression Inventory-II; PCL-5 – Posttraumatic Stress Disorder Checklist; SCID-5-RV <u>–</u> Structured Clinical Interview for DSM-5.

| *Variable* | **Anhedonia Group**<br>**(n=25)** |  |  |
| --- | --- | --- | --- |
|  | *M* | *SD* | *Range* |
| PSS | 20.84 | 3.64 | 13 - 27 |
| SHAPS | 36.64 | 4.37 | 30 - 45 |
| BDI-II Anhedonia Subscale | 5.04 | 2.03 | 2 - 9 |
| BDI-II Total | 20.20 | 9.12 | 3 - 41 |
| PCL-5 | 19.12 | 12.38 | 1 - 43 |
| Primary Diagnosis (SCID-5-RV) |  |  |  |
| *No Current Diagnosis* | 6 | (24%) |  |
| *Major Depressive Disorder (MDD)* | 9 | (36%) |  |
| *Persistent Depressive Disorder (PDD)* | 3 | (12%) |  |
| *Generalized Anxiety Disorder* | 3 | (12%) |  |
| *Attention-Deficit Hyperactivity Disorder (ADHD)* | 2 | (8%) |  |
| *Specific Phobia* | 1 | (4%) |  |
| *Other Specified Anxiety Disorder* | 1 | (4%) |  |

ANH and CON groups did not differ in age (*t*(28.7) = -0.14, *p* = .887). There were significantly fewer females in the CON group (χ^2^(1) = 6.13, *p* = .013). [^11^C]Raclopride dose differed between groups; for the ANH and CON groups, the average dose was 13.27 mCi (*SD* = 1.28) and 11.73 mCi (*SD* = 2.14), respectively (*t*(14.9) = 2.31, *p* = .036). Thus, analyses presented here controlled for sex and [11C]Raclopride dose.

ANH participants reported moderate levels of anhedonia, as assessed by the SHAPS, as well as moderate depressive symptoms, as assessed by the BDI-II (45). ANH participants’ PSS scores reflect moderate stress (46), and PCL-5 scores reflect mild stress (35). Five ANH participants had PCL-5 scores of 33 or greater, indicating clinically significant PTSD symptoms.

Within the ANH group, males reported significantly greater perceived stress on the PSS than females (*t*(22.7) = -2.73, *p* = .011). Anhedonia severity ratings did not differ by sex. In the ANH group, scores on the SHAPS and BDI-II anhedonia subscale were positively correlated (*r* = 0.65, *p* = .0005) and PSS and BDI-II anhedonia subscale scores were positively correlated (*r* = 0.47, *p* = .0179). Six ANH participants did not meet criteria for any current diagnoses; however, each had a CGI-S score of 3, indicating clinical impairment. See Supplemental Materials IIA for task reaction time and valence ratings analyses.

### Striatal Dopaminergic Functioning

#### Group Differences in ΔBP_ND_ during the MID Task (Reward – Neutral Conditions)

Striatal clusters in the left putamen, right putamen and pallidum, left caudate, and left nucleus accumbens (NAc), extending into the left putamen, demonstrated between-group differences in ΔBP_ND_ values (ANH - CON) for the Reward - Neutral contrast, *F*’s(1,20) > 7.38, *p*’s < .01. These analyses controlled for sex and [^11^C]raclopride dose, given that there were significantly fewer females in the CON group and [^11^C]raclopride dose was, on average, higher in the ANH group. See *Table 3* for striatal cluster statistics. *Figure 3* shows [^11^C]raclopride BP_ND_ values for each participant by condition and group. Relative to CON participants, ANH participants showed higher [^11^C]raclopride BP_ND_ during the reward condition relative to the neutral condition (*Figure 3*). This finding indicates that, relative to CON participants, ANH participants exhibited reduced task-related DA release to rewards in the striatum. Results for exploratory PET analyses are reported in Supplemental Materials IIB-IIE.

**Table 3.** Striatal Clusters demonstrating ANH - CON Group Differences in ΔBP_ND_ values (Reward - Neutral) at a threshold of z > 2.58 (uncorrected). Contrast of ANH - CON; Reward - Neutral ΔBP_ND_ values. MNI Coordinates. For all clusters, the Group (ANH, CON) × Condition (Reward, Neutral) interaction effect on [^11^C]raclopride ΔBP_ND_ values were significant, controlling for sex and [^11^C]raclopride dose. This was expected given that ANOVA results are dependent on the cluster-defining contrast. *p*-values <.05*, <.01**, <.001***. NAc, Nucleus Accumbens. ANH, Anhedonia participants. CON, Control participants.

| *Cluster Label* | *Cluster Size* | *Max Z value* | *Max X* | *Max Y* | *Max Z* | *Group x Condition Interaction p-value* |
| --- | --- | --- | --- | --- | --- | --- |
| Left putamen | 88 | 4.70 | -22 | 4 | 12 | .0006** |
| Right putamen/pallidum | 23 | 3.63 | 18 | 6 | -4 | .007** |
| Left caudate | 23 | 3.33 | -16 | 4 | 14 | .010* |
| Left NAc and putamen | 19 | 3.45 | -12 | 6 | -8 | .009* |

**Figure 3.**
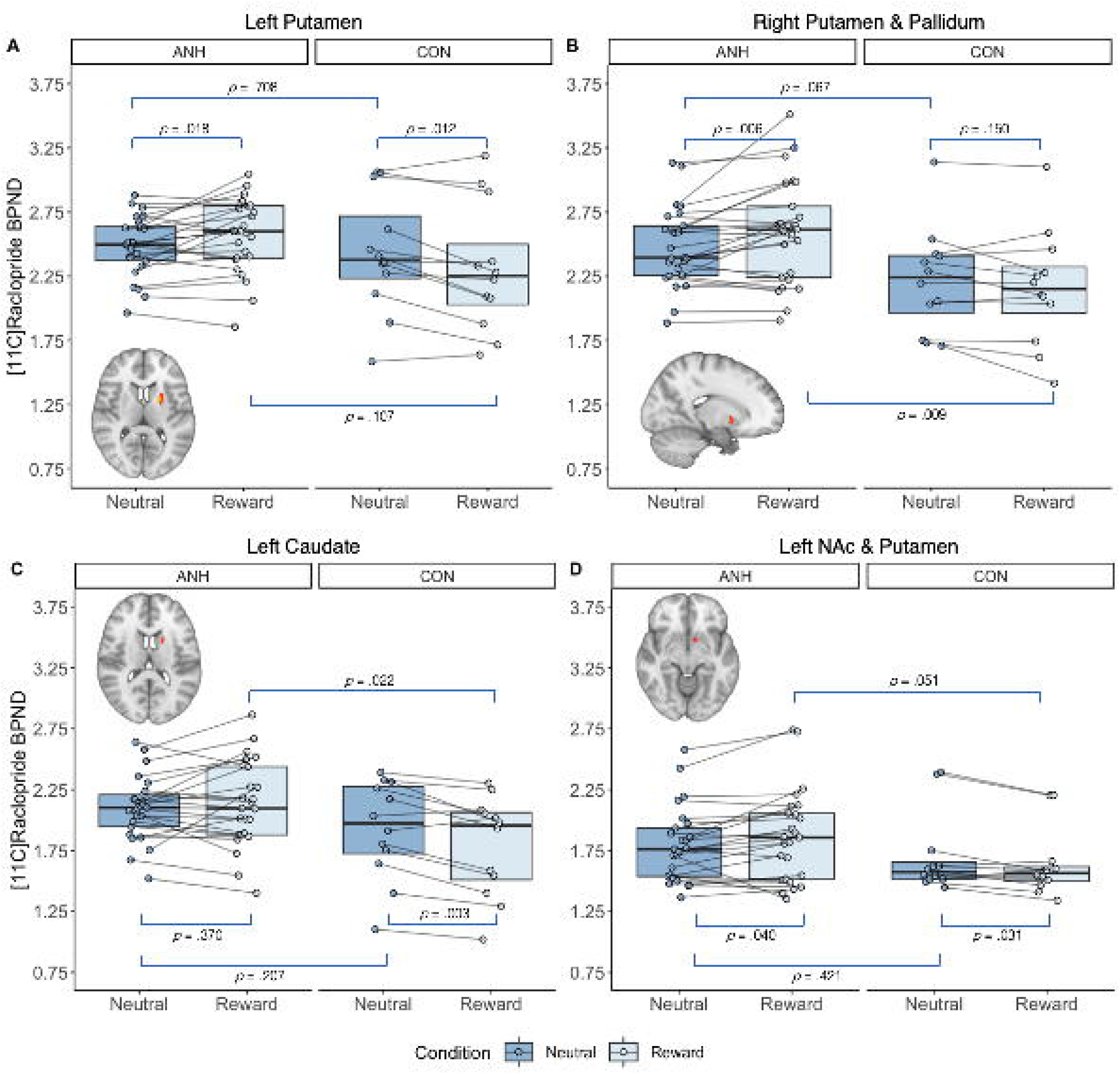
[^11^C]Raclopride binding potential in functionally-defined striatal clusters demonstrating group differences for the contrast of (ANH - CON; Reward - Neutral). T-tests shown here (blue lines) are within-group comparisons of ΔBP_ND_ values (Reward - Neutral) and between-group comparisons of BP_ND_ values (Neutral). In each of these four clusters, there was a significant group × condition interaction, Fs(1,20) > 7.38, ps < .010. This was expected given that ANOVA results are dependent on the cluster-defining contrast. The neutral phase depicted here encompasses the first 42 minutes of scanning (i.e., a measure of baseline DA).

### fMRI Activation

Results of fMRI activation analyses are presented in Supplemental Materials IIF. These analyses showed a single between-group difference in right caudate activation, at an uncorrected threshold, which was not associated with clinical measures of anhedonia or self-reported stress.

### fMRI Connectivity

#### PET-derived Seed-based General Functional Connectivity

Whole-brain GFC analysis revealed several significant group differences. PET-derived seeds demonstrated negative connectivity with subcortical and cortical regions in the ANH group, relative to the CON group. Target regions of these seeds included structures commonly implicated in reward processing, including bilateral caudate, putamen, and pallidum, as well as the medial prefrontal cortex. Associated regions in the anterior cingulate cortex and the thalamus were also identified as target regions. See *Table 4* for connectivity statistics. *Figure 4* illustrates group differences in GFC between the PET-derived seeds and their respective target regions.

**Table 4.** Statistics for clusters demonstrating ANH - CON group difference in GFC seed-to-voxel analysis with PET-derived seeds. Size p-values indicate the significance of the size of the target cluster (voxels). Peak p-values indicate the significance of the signal of the target cluster, at its peak, or strongest point of connectivity. FWE, family-wise error. FDR, false-discovery rate. Unc, uncorrected. FWE and FDR are two common methods for correction of multiple comparisons. Unc p-values have not been corrected for multiple comparisons. ANH, Anhedonia participants. CON, Control participants.

| Seed<br>*Target Label* | Cluster<br>Size<br>(voxels) | Size<br>*p*-FWE | Size<br>*p*-FDR | Size<br>*p*-unc | Peak<br>*p*-FWE | Peak<br>*p*-unc |
| --- | --- | --- | --- | --- | --- | --- |
| Left Putamen |  |  |  |  |  |  |
| *Bilateral Striatum*<br>(-22, 0, 6) | 786 | .000 | .000 | .000 | .001 | .000 |
| *Right Striatum*<br>(18, 6, 8) | 603 | .000 | .000 | .000 | .009 | .000 |
| *Right Superior Frontal Gyrus*<br>(22, -4, 62) | 87 | .010 | .006 | .000 | .997 | .000 |
| Right Putamen / Pallidum |  |  |  |  |  |  |
| *Right Striatum*<br>(18, 8, -8) | 268 | .000 | .000 | .000 | .003 | .000 |
| *Right Paracingulate Gyrus /*<br>*Anterior Cingulate Gyrus*<br>(2, 36, 26) | 78 | .014 | .008 | .000 | .159 | .000 |
| *Right Caudate*<br>(12, 6, 12) | 54 | .085 | .033 | .001 | .975 | .000 |
| Left Caudate |  |  |  |  |  |  |
| *Bilateral Striatum / Left Thalamus*<br>(18, 18, -4) | 515 | .000 | .000 | .000 | .397 | .000 |
| *Left Caudate*<br>(-16, 10, 20) | 324 | .000 | .000 | .000 | .001 | .000 |
| *Left Striatum*<br>(-24, 2, -12) | 99 | .005 | .002 | .000 | .987 | .000 |
| *Left Caudate / Thalamus*<br>(-10, -12, 16) | 69 | .035 | .011 | .000 | .148 | .000 |
| Left Nucleus Accumbens and Putamen |  |  |  |  |  |  |
| *Left Striatum*<br>(-14 6, -12) | 268 | .000 | .000 | .000 | .003 | .000 |
| *Medial Frontal Cortex*<br>(-8, 50, -16) | 57 | .065 | .038 | .001 | .978 | .000 |

**Figure 4.**
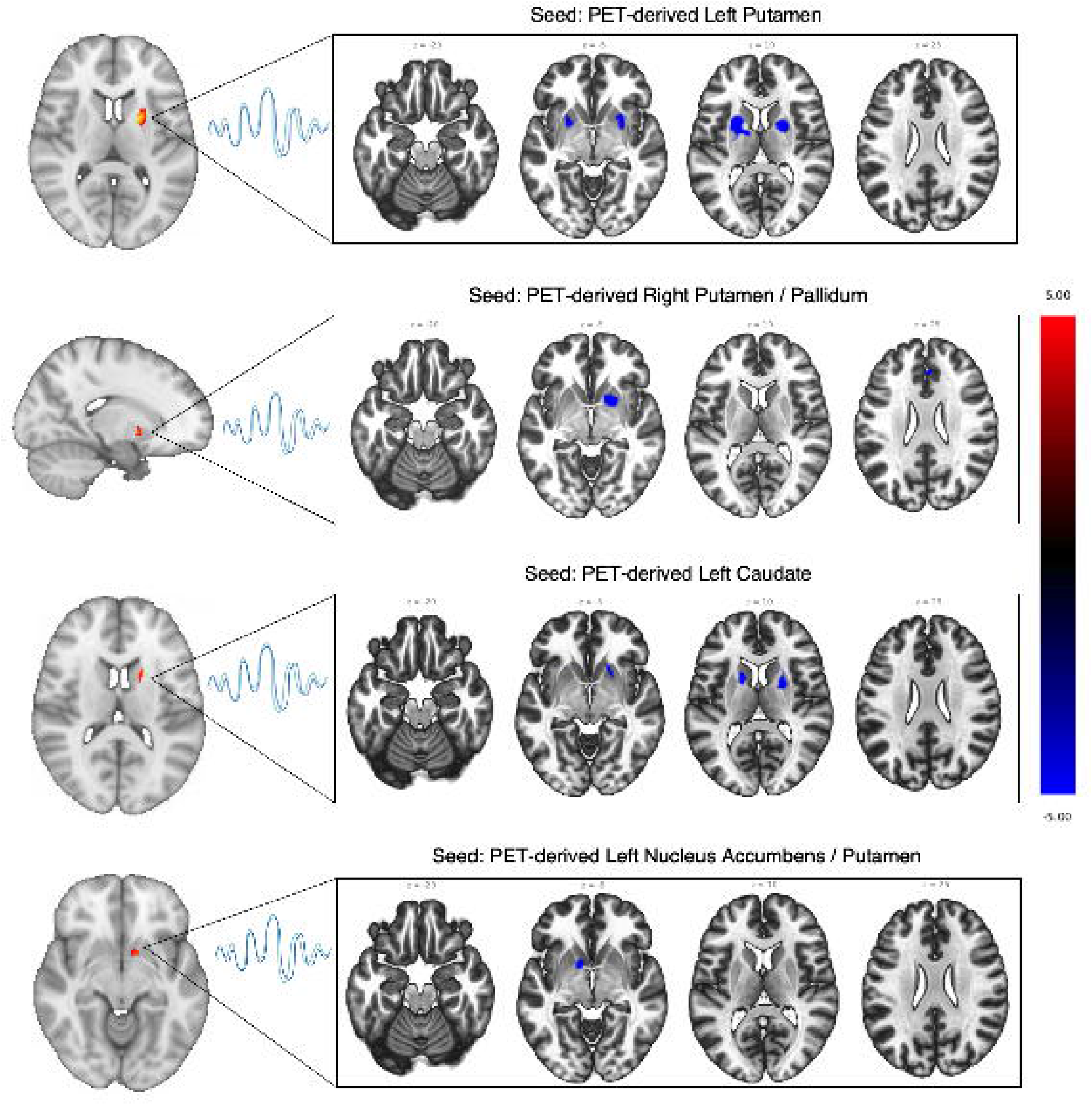
Group differences in general functional connectivity of PET-derived seeds. Seed-to-voxel analysis (ANH - CON) controlling for age and sex. Only negative connectivity values were found in the ANH group, represented in blue. PET striatal seeds are presented in radiologic view, so the left and right are reversed. ANH, Anhedonia participants. CON, Control participants.

### Relations between Anhedonia and Mesocorticolimbic Network Functioning

#### Anhedonia and Task-Related DA Release in Functionally-defined Striatal Clusters (PET)

In the ANH group, we examined associations between ΔBP_ND_ values in the above striatal clusters that demonstrated group differences and anhedonia severity scores on the SHAPS and BDI-II anhedonia subscale. Reduced task-related DA release to rewards in the left putamen cluster was significantly associated with BDI-II anhedonia scores (β_*STD*_ = .47, *SE* = 0.18, *t* = 2.57, *p* = .017, *p*_FDR_ = .137) (*Figure 5*). BDI-II anhedonia subscale scores were not significantly associated with task-related DA release in the other three striatal clusters (*p*’s > .05). SHAPS scores were not significantly associated with task-related DA release in any of the four striatal clusters (*p*’s > .05). Results for exploratory analyses with anhedonia severity are reported in the Supplemental Materials IIG-IIH.

**Figure 5.**
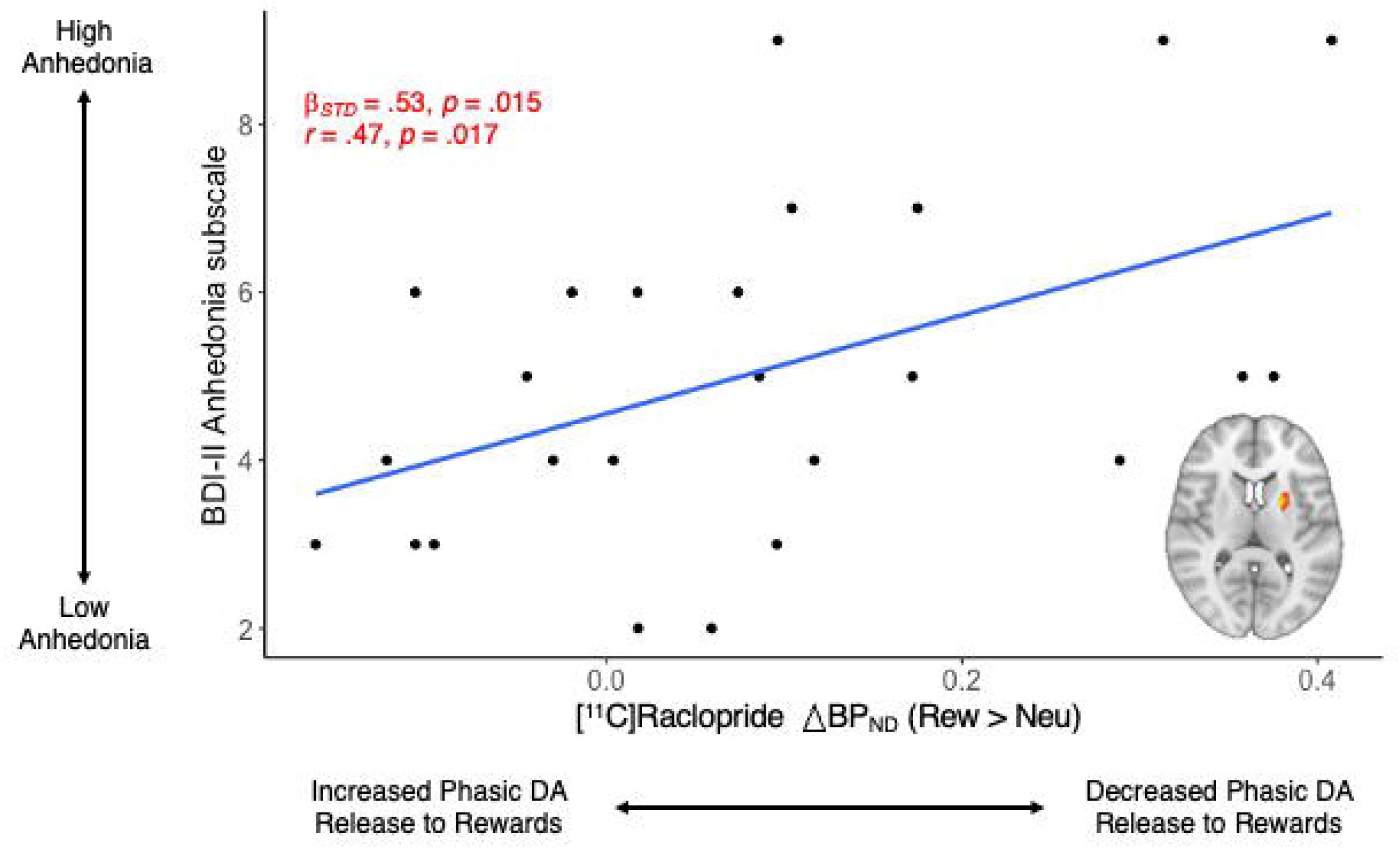
Task-related DA release to rewards in the functionally-defined left putamen striatal cluster predicts BDI-II anhedonia subscale scores for ANH participants (n=25). In ANH participants, greater [^11^C]raclopride DBP_ND_ was associated with greater anhedonia severity on the BDI-II anhedonia subscale. Positive DBP_ND_ values represent decreased task-related DA release to rewards, relative to neutral stimuli, on the MID task. BDI-II, Beck Depression Inventory.

#### Anhedonia and Mesocorticolimbic Network Connectivity (PET-MR)

Neither the SHAPS nor BDI-II anhedonia subscale were significantly associated with PET-derived GFC strength for any region pairs (*p’s* > .05) (see *Figure 6*).

**Figure 6.**
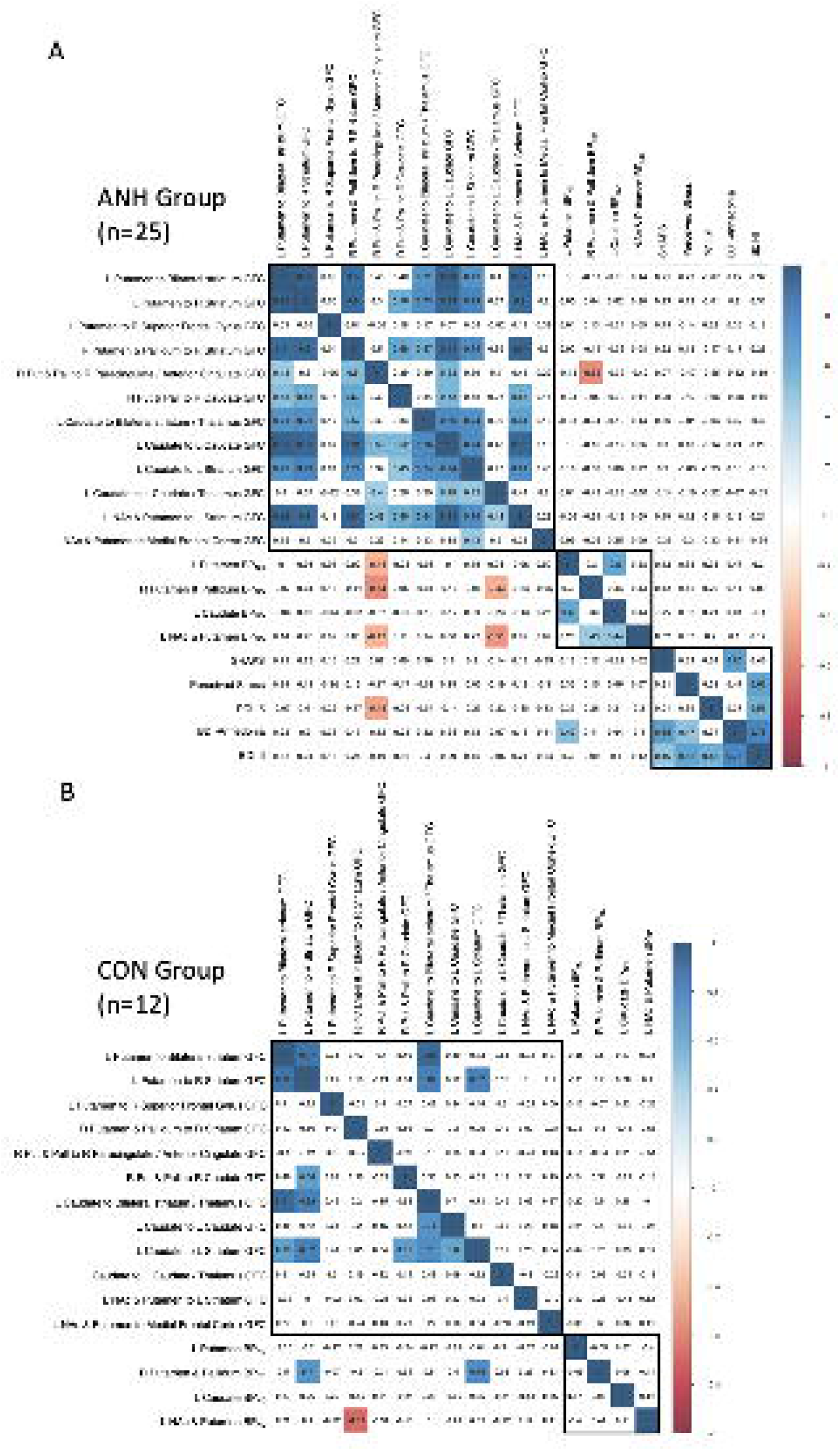
Pearson correlation matrix for variables of interest. Pearson correlation values range from -1 to 1. Significant correlations (*p* < .05) are displayed in color; non-significant correlations (*p* > .05) are displayed in white. Correlations presented in the upper right triangles of the matrices are corrected for multiple comparisons, using the false-discovery rate (FDR) method. Correlations in the lower left triangles are uncorrected. L, left. R, right. GFC, general functional connectivity. Put, Putamen. Pall, Pallidum. NAc, Nucleus Accumbens. ΔBP_ND_, binding potential non-displacement. SHAPS, Snaith-Hamilton Pleasure Scale. PCL-5, PTSD Checklist for DSM-5. BDI, Beck Depression Inventory. ANH, Anhedonia. CON, Control.

### Relations between Self-Reported Stress and Mesocorticolimbic Network Functioning

#### Self-Reported Stress and Task-related DA Release to Rewards in Functionally-defined Striatal ROIs (PET)

Within the ANH group, analyses with the PSS and PCL-5 yielded no significant associations between self-reported stress and mesocorticolimbic task-related DA release to rewards in striatal clusters. Results for exploratory analyses with self-reported stress are reported in Supplemental Materials III-IIJ.

#### Self-Reported Stress and Mesocorticolimbic Network Connectivity (PET-MR)

Exploratory analyses with the PCL-5 yielded one significant association with GFC between the PET-derived right putamen and pallidum cluster and a target region in the paracingulate and anterior cingulate cortex; however, this association between PCL-5 scores and GFC was not significant after FDR-correction for multiple comparisons. Scores on the PSS were not significantly associated with mesocorticolimbic network connectivity.

### Correlations between [^11^C]Raclopride Binding Potential, fMRI Network Connectivity, and Clinical Measures

Figure 6 summarizes bivariate Pearson correlations in the ANH group between primary and secondary clinical measures of stress and anhedonia, [^11^C]raclopride binding potential in striatal clusters demonstrating group differences, and general functional connectivity of these striatal clusters with their respective whole-brain target regions. As hypothesized, greater [^11^C]raclopride binding potential (i.e., reduced striatal DA release to rewards) in striatal clusters tended to be negatively associated with general functional connectivity values of these seeds and their target regions (see *Figure 6, orange boxes in lower triangle*). However, not all of these correlations remained after an FDR-correction for multiple comparisons (see *Figure 6, upper right triangle*).

## Discussion

This investigation explored associations among anhedonia, striatal DA binding, and reward circuitry functioning in a transdiagnostic sample with clinically impairing anhedonia. Stress was also examined in an exploratory manner.

### Striatal Dopamine and Anhedonia

Extending previous findings of decreased striatal DA release to rewards in MDD (8,17), we found reduced striatal DA release to rewards in ANH participants. Interestingly, in the ANH participant group alone, there were no regions that showed a significant change in ΔBP_ND_ from the neutral to reward condition of the MID. This null finding suggests that participants with anhedonia demonstrate a blunted DA response to rewards. Next, we found that relative to the CON group, ANH participants exhibited increased [^11^C]raclopride ΔBP_ND_ in the left and right dorsal striatum and left ventral striatum (*Figure 3*). Together, these findings represent the first report of reduced task-related DA release to rewards in a transdiagnostic sample with clinically impairing anhedonia.

Reduced striatal DA release to rewards in ANH participants may reflect impaired reward learning (1) although this was not seen behaviorally. The optimized MID task used here required learning which cues predicted differing reward magnitudes, enhancing the sensitivity of the task to positive prediction errors encoded by task-related DA release (4). Though we did not evaluate prediction errors *per se*, impaired modulation of behavior by rewards during a probabilistic reward task is characteristic of anhedonia in individuals with MDD (47,48). Although Hamilton and colleagues (2018) also reported lower availability of DA during baseline in an MDD sample, our findings are not consistent with this interpretation in the ANH group (17). That is, we did not find evidence that ANH participants were characterized by significantly lower baseline DA relative to CON participants (see *Figure 3, for comparisons of neutral phase BP*_*ND*_). Supplemental results for baseline differences in raclopride binding potential during the neutral condition, using an ROI approach, are presented in Supplemental Materials IIE.

Regarding associations between striatal ΔBP_ND_ and anhedonia, we found that increased ΔBP_ND_ in the left putamen, indicative of decreased task-related DA release, was positively associated with anhedonia severity on the BDI-II anhedonia subscale. Within functionally-defined striatal clusters, SHAPS scores were not significantly related to task-related DA reward signaling, which is consistent with at least one [^11^C]raclopride PET study in MDD (8). These contrasting results between the BDI-II anhedonia subscale and the SHAPS may be due to differences in the aspects of anhedonia that these two scales capture. Whereas the SHAPS primarily assesses aspects of consummatory reward capacity (i.e., pleasure) (49,50), the BDI-II anhedonia subscale captures aspects of both consummatory and anticipatory (i.e., motivation or interest) reward capacity (37,50,51). Nevertheless, our finding of an association between striatal ΔBP_ND_ and anhedonia on the BDI-II anhedonia subscale requires replication, particularly because the internal consistency of the BDI-II anhedonia subscale is not outstanding (α = 0.60). The internal consistency of the SHAPS has been shown to be higher than the BDI-II anhedonia subscale (52).

### fMRI Activation during Reward Anticipation and Reward Outcome

We did not find evidence of altered mesocorticolimbic activation during reward anticipation or reward outcome phases in ANH participants after correcting for multiple comparisons (see *Supplemental Materials IIF*). The lack of fMRI activation differences may be attributable, in part, to inadequate power of the current study to detect smaller effects given the small sample. Prior fMRI research has shown hypo-responsivity of striatal regions during anticipatory (1,53,54) and consummatory processing (55–57) in psychiatric populations where anhedonia is a central feature. As stated above, the optimized MID task used here requires learning whereas the standard fMRI MID task does not, and this could contribute to differences relative to prior MID studies. Given that DA is important for learning, the current design is an improvement with reference to DA. The current study’s uncorrected fMRI activation results should be cautiously considered within the broader literature. Additionally, recent PET-MR investigations of striatal DA binding in MDD did not report group differences in fMRI activation during reward anticipation or reward outcomes (17,19).

### Anhedonia and Mesocorticolimbic General Functional Connectivity

The present study also investigated functional connectivity seeded by regions exhibiting blunted striatal DA release to rewards (i.e., PET-derived seeds) using a whole-brain GFC approach. Compared to CON participants, ANH participants showed negative GFC between PET-derived seeds and several regions implicated in reward processing (i.e., bilateral caudate, putamen, and pallidum), as well as cognitive control (e.g., anterior cingulate gyrus) and control of attention (e.g., thalamus). These results are consistent with reports of altered functional cortico-striatal connectivity in MDD (12,58,59) and a previous [^11^C]raclopride PET-MR study of functional connectivity in MDD (17). In MDD, increased ΔBP_ND_ in the ventral striatum predicted decreased functional connectivity between PET-derived seeds and default-mode and salience network regions (17).

### Impact of Stress on Anhedonia via Striatal Dopamine

Stress is believed to desensitize the mesocorticolimbic DA system and contribute to the maintenance of anhedonic behavior (3,26,60). We hypothesized that self-reported stress on the PSS would predict anhedonia severity and be associated with striatal DA release to rewards, illustrating one potential mechanism linking self-reported stress and anhedonia. This was an exploratory hypothesis, given that the current sample was not selected for their exposure to stress. Consistent with previous work (3,61), perceived stress and scores on the BDI-II anhedonia subscale were significantly correlated (*r*=.47). However, we did not find evidence for the contribution of self-reported stress on mesocorticolimbic DA system functioning. The PSS, our primary measure of self-reported stress, is a retrospective measure that assesses the extent to which stress is unpredictable and uncontrollable during the last month, but it does not assess different types and chronicity of stressors (62). It is possible that other scales that objectively assess stressful life conditions and situations may be better suited to illuminate the role of self-reported stress in DA function. Furthermore, the nucleus accumbens (NAc) is strongly implicated in stress regulation (20) and demonstrates a blunted response during reward consumption in patients with MDD (63). Here, group differences in dopaminergic response to rewards (*Figure 3*) highlighted one small cluster (size=19 voxels) located between the left NAc and left putamen. We may not have found evidence of a relation between self-reported stress and mesocorticolimbic DA system functioning because these clusters were primarily located outside of the NAc.

### Limitations and Future Directions

This study has a number of limitations. First, the sample size, though comparable to many PET studies (64), was modest. As such, the PET analyses presented are not corrected for multiple comparisons. Future work that attempts to replicate this study in larger samples should aim to use a more stringent method of statistical correction. Second, given that this is a cross-sectional study, we cannot determine causal relationships between reduced striatal DA release to rewards and anhedonia. Future research should investigate temporal relations between reduced striatal DA release to rewards and anhedonia. Third, the ANH sample was not recruited based on severity of self-reported stress. Although the ANH sample demonstrated moderate levels of stress (see *Table 2*), the mean level of PSS scores was lower than psychiatric samples and the variability of PSS scores was limited (65). Relatedly, the current study sought to examine how stress, broadly measured, relates to striatal reward responses. While we did not find any effects with respect to self-reported stress, exploring DA reward signaling during a PET-MR stress-related paradigm would be one area for future study. Fourth, while the current study did not find any dependence on total injected dose, it is possible that other unknown factors could affect the dynamic uptake in the studied regions and the reference region differently.

In summary, the present study is the first to investigate task-related striatal DA release to rewards in a transdiagnostic anhedonic sample. This study provides support for the association between blunted striatal DA functioning and transdiagnostic anhedonia. We found blunted general functional connectivity with PET-derived striatal seeds in anhedonia participants, but these group differences were not associated with anhedonia severity. We demonstrated that self-reported stress was strongly associated with anhedonia but was not associated with striatal DA binding. These findings provide support for the association between stress and anhedonia and highlight a potential molecular mechanism of impaired reward processing in anhedonia-related psychopathologies.

## Data Availability

All data produced in the present study are available upon reasonable request to the authors

## Acknowledgments

This research was supported by R61/R33 MH110027 to GSD and MJS, R21 MH110933 to GSD and JMH, K23 MH113733 to ECW, and UL 1TR002489. DAP was partially supported by R37 MH068376 and R01 MH095809. DD was partially supported by R01 MH111676. The content is solely the responsibility of the author and does not necessarily represent the official views of NIH.

