## Supplemental Materials for "Striatal Dopamine Binding in Anhedonia: A Simultaneous [^11^C]Raclopride Positron Emission Tomography and Functional Magnetic Resonance Imaging Investigation"

I. **Methods**
A. Eligibility Criteria

Individuals who met any of the following criteria were excluded from participation: (1) disorders whose medication management is the primary treatment approach (i.e., bipolar disorder or mania, schizophrenia, or other psychotic disorders), (2) prior treatment with behavioral activation or mindfulness-based approaches for depression (i.e., prior exposure to the experimental treatments used in the parent study), (3) difficulty understanding the cognitive components of treatment (i.e., an intellectual disability, neurocognitive disorder, dissociative disorder, or IQ score less than 90), (4) having a feeding or eating disorder which may have confounding effects on the BOLD fMRI signal, (5) severe current or lifetime substance use disorder (SUD) or alcohol use disorder (AUD) which may have confounding effects on the BOLD fMRI signal, (6) current suicidal intent or plan within the last month (i.e., those recommended for referral to more intensive clinical management services), (7) psychotropic medication use within the last month and/or current psychotherapy, (8) currently pregnant (i.e. measured via urine pregnancy screen immediately before MRI scans), (9) positive urinalysis screen for substance use at the time of the MRI scan, (10) neurological conditions (i.e., history of stroke, seizure, or traumatic brain injury), (11) contraindications for MRI imaging (i.e., metal in the body, prior metallic injury, or metallic dental work, (12) PET scans in the prior 12 months that exceed UNC IRB guidelines of 15 mSv radiation exposure, (13) radiation therapy or chemotherapy in the 2 months prior to scanning, and (14) blind or unable to read and comprehend English.

B. PET Processing

List mode 3-D emission data were collected starting from bolus injection (beginning approximately 1 min after the scan began) and continued over the 75 min scan. PET acquisition took place for 63 minutes. Radioactivity was limited to 15mCi in total over the bolus and infusion and mass dose did not exceed 10µg for the duration of the scan. In the first portion of scan acquisition, after the PET scan was initiated, participants underwent two 8-min fMRI resting state scans and one 6-min high resolution T1 scan. In the second portion, participants completed the monetary incentive delay (MID) task described below, during which BOLD fMRI data were acquired simultaneously. A structural T1 MR sequence (FOV=256 mm, 111 mm resolution, TR=2530ms, TE=1.69ms, flip angle=7 degrees) was used for anatomical localization, spatial normalization of imaging data, and generation of attenuation correction maps. Two identical resting-state scan sequences (echo planar imaging, FOV=212 mm, 3.312 x 3.312 x 3.3 mm resolution, TR=3000, TE=30ms, flip angle=90 degrees) were obtained to capture endogenous neural activity. The functional scan sequence (same parameters as resting-state), during which participants were engaged in the MID task, was collected over three task blocks.

**Reconstruction.** Attenuation maps including bone and sinus detail were created using the PseudoCT method (1), which uses the subject’s Dixon attenuation map and the T1 MPRAGE image to estimate a CT- equivalent attenuation map. The PseudoCT method showed high accuracy in a head-to-head comparison of several methods for MR-based attenuation correction 2. PET images were reconstructed from list-mode data for 70 1-minute time frames starting at injection time using vendor-provided software (E7Tools, Siemens Healthineers). Reconstructions used the OS-EM algorithm for 3 iterations and 24 subsets and included corrections for attenuation, scatter, and randoms. The reconstruction grid was 344×344 with 127 axial slices and a voxel size of 2.086mm × 2.086mm × 2.032mm.

**Motion Correction.** The dynamic PET images were corrected for motion using the Realign procedure of SPM12 3. This method computes a rigid transformation for each time frame to align all to a common reference. The result of this procedure is a set of 70 × 1-minute dynamic images that are well aligned. As a quality-control measure, the motion-corrected frames were observed in cine mode to detect possible errors. In all but two cases, the motion correction was found to achieve good alignment. For one subject we found a significant sudden movement over three frames that was not well corrected. Because this only affected three frames in the early portion of the scan, we chose to exclude those frames from the dynamic fits for that subject alone. In another subject, movement during the scan was found to be so substantial and uncorrectable that the subject was excluded from further analysis.

**Subject-to-MNI Space Transformation.** For each subject, a mapping from subject space to reference MNI space was determined as a composition of two transforms: first, a rigid transform to align the motion-corrected PET to the subject’s T1 MPRAGE image, and, second, a deformable transform to align the subject’s T1 MPRAGE image with the MNI reference T1 image. The rigid transform was found to be necessary in many subjects either due to (1) movement between the times that PET and T1 images were taken and/or (2) the motion correction procedure displacing the PET from the T1 by a few millimeters. The rigid transforms were determined manually by displaying the motion- corrected PET (20-minutes post-injection) overlaid on the T1 MRI in Slicer 4. The deformable component was determined using the Dartel tools in SPM12 5. This procedure solved for a mean intermediate reference among all subjects and the MNI reference based on segmentations of gray and white matter from the T1 MPRAGE images. Then, deformations for the individual subjects mapping to the MNI space were composed from subject- >intermediate and inverse (intermediate->MNI) deformations. The result of this procedure was a set of rigid (PET->T1) and deformable (T1->MNI) transforms for each subject.

**Subject-specific Atlases.** With the individual transformations available, atlases defined in the MNI space were mapped to individual subject PET spaces by inverting the transforms determined in part C above. The AAL3 atlas 5, which is defined in 2mm MNI space, was transformed to each subject’s PET space and resampled to the 344×344×127 PET grid so that the atlas could be applied to each of the 70 motion-corrected PET images without further transformation. The AAL3 atlas was mapped using nearest-neighbor interpolation to preserve the integer labeling of its 170 regions. In addition, the three-region 1mm striatum structural atlas 6, also defined in MNI space, was divided by left and right hemispheres (making six total regions), then transformed in the same way; however, this atlas was mapped using linear interpolation in order to weight edge voxels for partial volume effects.

**BP_ND_ Estimates from Regional TACs.** For each subject, the subject-specific 170-region AAL3 atlas was applied to each of the 70 motion-corrected PET images to obtain the mean PET intensity (in Bq/ml) in each region for each time frame. Thus, a time-activity curve (TAC) was obtained for each region. The simplified reference tissue (SRTM) model 7 was applied to TAC data as follows. The SRTM model was applied with a two-part BP_ND_ component to account for the neutral (from injection time to start of reward task) and reward states:

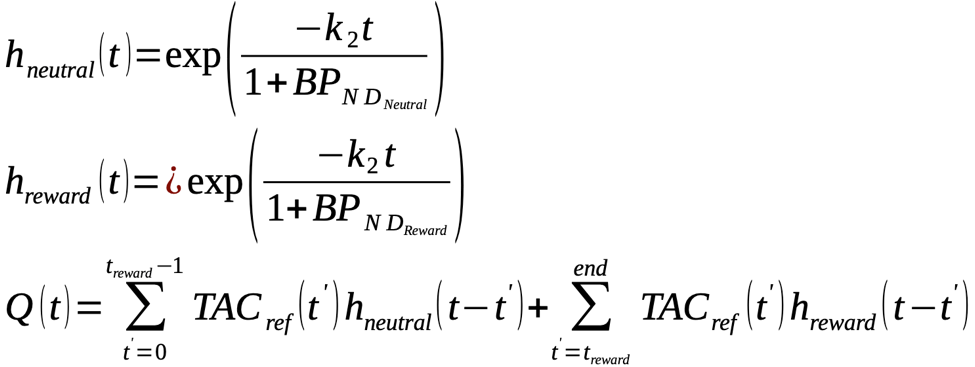

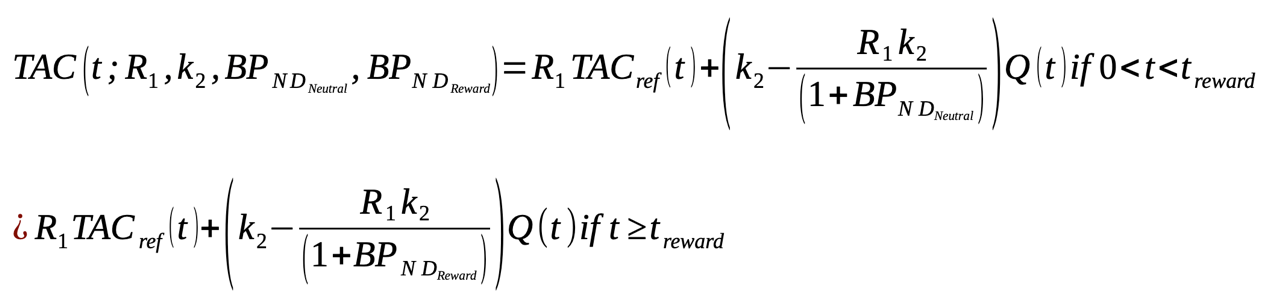

Where

- *t*=0 represents the start of the scan at time of bolus injection
- *h_neutral_(t)* and *h_reward_(t)* are the exponential system impulse responses in the reward and neutral states, respectively;
- *Q (t)* is a time-dependent discrete convolution of the reference TAC with the system response kernels accounting for the reward and neutral conditions;
- *TAC (t)* represents the TAC of a given atlas region;
- *TAC_ref_ (t)* represents the TAC for the cerebellar reference region (measured from the cerebellar regions of the AAL3 atlas, excluding regions labeled as vermis);
- *R*_1_ is an estimated parameter of the SRTM model representing the ratio of kinetic transport rates from plasma to free-tracer tissue compartments in the TAC region studied and reference region;
- *k_2_* is an estimated parameter of the SRTM model representing the kinetic transport rate from free-tracer tissue to plasma compartments in the TAC region studied;
- *BP_NDNeutral_* and *BP_NDReward_* are estimated parameters representing the non-displaceable binding potential in each of the two task states, Neutral and Reward, respectively;
- *t_reward_* is the time at which the reward task is begun (usually 42 minutes into the study but varied for individual subjects based on the recorded task start time; the baseline binding potential is estimated from injection up to start of reward block).

For each TAC, the two-part model was fitted with a custom MATLAB script applying a nonlinear least-squares fit to the SRTM model. Thus, for each subject and for each hypothesized atlas region, we obtained estimates of BP_ND_Neutral_ and BP_ND_Reward_. For regional estimates from the six-region striatal atlas, the cerebellum reference TAC from the AAL3 atlas was used as the reference TAC.

**BP_ND_ Voxel Maps.** To perform voxel-wise analysis in the common MNI space, a set of BP_ND_ maps was created for each subject. For each subject, the 70 motion-corrected PET images were smoothed with an 8mm Gaussian kernel to reduce noise for individual voxel fits. A mask was applied to screen out voxels with little PET activity. Then, a TAC was obtained for each voxel over the 70 time frames, and the two-part SRTM model defined in part E was applied to obtain BP_ND_Neutral_ and BP_ND_Reward_ for each voxel within the brain mask. This resulted in subject-space maps of BP_ND_Neutral_ and BP_ND_Reward_. A map of fit quality in each voxel, based on the coefficient of determination from each fit, was also produced and checked as a quality-control procedure. There were no issues observed with fit quality in the striatal regions.

The subject-space maps were then transformed to the MNI space using the transformations of part C. This allowed the individual subject maps to be compiled in the common MNI space, creating maps of mean and variance of BP_ND_R-N_ (Reward-Neutral) for each group (Control and ASD) and corresponding z-score maps.

C. PET-MR Monetary Incentive Delay (MID) Task

Participants completed a monetary incentive delay (MID) task, which has been shown to reliably elicit corticostriatal BOLD responses (2) and phasic DA release to rewards (3). The MID task, presented using PsychoPy software version 1.84.1 (4) was optimized for the slow kinetics of [^11^C]raclopride displacement (5) and has been used in prior investigations by our research team (6). This optimized MID task includes novel features designed to maximize detection of DA release in the PET-MR environment. First, the initial reward block begins approximately 40 min after the [^11^C]raclopride bolus injection, after the target-to-reference region ratio is stabilized. This long uptake period serves as a baseline scan. Second, about 75% of reward trials are followed by reward feedback, resulting in a success rate that is higher than traditional MID tasks to enhance incentive motivation. Third, while most MID versions use explicit reward and neutral cues that make the potential outcome of each trial clear, the current design forces participants to learn which cues predict which reward magnitudes. By adding associative learning, the current design aims to enhance sensitivity to positive prediction errors (and other learning-related signals) encoded by phasic DA release (7). This modified version of the MID task (*Figure 2)* was developed at McLean Hospital (Drs. Dillon and Pizzagalli).

During the neutral block, participants completed 63 trials that started with a square cue. No monetary rewards were delivered on these trials. Instead, sufficiently speeded button presses resulted in the presentation of a gray rectangle as a “no-reward” outcome. The other outcomes indicated either no response (“No Response!”), the response was too quick (within 100 ms of the target presentation: “Too Fast!”), or it was made after an adaptive reaction time (RT) threshold (“Too Slow!”) that was programmed such that ~75% of each participant’s responses were successful.

In the reward blocks, comprised of 75% rewarded and 25% nonrewarded trials, participants won money if they responded quickly enough to the target stimuli on rewarded trials. In the reward blocks, different polygon cues (square, triangle, pentagon, and hexagon) indicated that trials could result in no-reward (gray rectangle) or a small (50 cents), medium (1 dollar), or large reward (5 dollars), respectively. The assignment of the four polygons to the four outcomes was stable across the reward blocks and counterbalanced across participants. Successful trials (i.e., trials with sufficiently speeded button presses) ended with images depicting the no-reward, small, medium, and large reward outcomes. Unsuccessful trials yielded the same feedback as in the neutral block (“Too Fast!”, “Too Slow!”, or “No Response!”). Each reward block contained two reward runs (number of trials per reward run: Block 1: 34/33, Block 2: 33/34). Following each neutral and reward block, participants rated cues and outcomes using a nine-point Likert scale with anchors of “very negative” and “very positive” at the ends and “neutral” in the center.

### D. fMRI Image Preprocessing & Motion

Freesurfer version 7.1.0 was used to reconstruct and remove non-brain tissue from anatomical (T1) scans. Functional data (i.e., MID task runs) for activation analyses were preprocessed using FSL FEAT version 6.0 (Oxford Centre for Functional Magnetic Resonance Imaging of the Brain (FMRIB), Oxford University, U.K.). The first four volumes of each functional run were discarded to allow for steady state equilibrium. Preprocessing in FSL included BET brain extraction for non-brain removal using the brain extraction tool (BET), motion correction using MCFLIRT, interleaved slice timing correction, spatial smoothing using a Gaussian kernel of FWHM 6mm, pre-whitening with the FILM tool (8), co-registration of functional and anatomical images using the boundary based registration (BBR) algorithm (9), registration to a standard stereotaxic space (Montreal Neurological Institute; MNI152 2mm) using linear transformation (FLIRT; 12 DOF, 10mm warp field resolution), and high-pass filtering (cutoff of 100 sec). To control for excessive motion, we censored volumes that exceeded a framewise displacement threshold of 0.5mm (10).

Functional connectivity data (i.e., resting-state and MID task runs) were preprocessed with the default preprocessing pipeline in the SPM12 CONN functional connectivity toolbox, version 19c. The default preprocessing pipeline consists of (a) resampling the scan data to 2 x 2 x 2-mm isotropic voxels and unwarping, (b) centering, (c) slice time correction, (d) normalization to standard MNI space, (e) outlier detection (ART-based scrubbing), and (f) spatial smoothing with an 8mm Gaussian kernel. Motion parameters were entered as multiple regressors and used to identify potential outliers that exceeded framewise displacement thresholds. For most of the sample (n = 35), all ﬁve runs of functional data were analyzable and all participants had at least three analyzable runs. Two subjects had excluded runs due to technical errors and striation artifacts. There were no signiﬁcant differences between groups on average motion, *t*(35) = -0.86, *p* = 0.396, or average global BOLD signal changes, *t*(35) = 0.78, *p* = 0.441.

E. Structural PET Analysis

In addition to functionally-defined clusters (see Main Text), we estimated BPND values for structural atlas-defined structural regions of interest (ROIs) in the striatum (i.e., bilateral caudate, putamen, and nucleus accumbens) using the Automated Anatomical Labelling (AAL3) atlas. For each significant atlas-defined cluster, t-tests were conducted to analyze group differences (ANH, CON) in the absolute differences (Reward-Neutral) of BP_ND_ values and Neutral BP_ND_ values. For clusters that showed between-group differences, we then examined associations between striatal BPND values and anhedonia and stress measures within the ANH group.

F. fMRI Activation Analysis

To examine fMRI responses during reward anticipation, the contrast between neutral and reward trials of all magnitudes (small, medium, and large) from the onset of the cue to the end of the fixation period (i.e., during the cue and the target) was examined. To examine fMRI responses during reward outcomes, the contrast between successful and unsuccessful outcomes (i.e., successful vs. unsuccessful reward outcomes on reward trials of all magnitudes (small, medium, and large)) was examined.

*A priori* hypothesis testing was conducted using a region of interest (ROI) approach. During reward anticipation, ROIs included the bilateral nucleus accumbens, caudate, and putamen (11,12). During reward outcomes, ROIs included the medial prefrontal cortex and anterior cingulate cortex (13). These ROIs were defined using the Harvard-Oxford subcortical and cortical structural probabilistic atlases. For each participant and condition, BOLD percent-signal change values were calculated and extracted from ROIs using FSL Featquery. We then conducted independent samples (ANH, CON) t-tests to explore group differences in BOLD percent-signal change. ROI analyses were supplemented with a general linear model approach, fitted to generate whole-brain images, allowing us to examine activation in other reward processing regions. Group-wise activation images were calculated using Bayesian estimation (FMRIB Local Analysis of Mixed Effects), and were cluster corrected with a cluster-defining threshold of z = 2.58, and cluster p-threshold of p < 0.05.

**II. Results**

A. Task Reaction Time and Valence Ratings

A Group (ANH, Control) × Cue (neutral, small reward, medium reward, large reward) ANOVA on reaction times revealed no main effect of Group or Group × Cue interaction, p’s > 0.44, but a main effect of Cue, F(3,105)=38.9, p<0.01. Follow-up t-tests examining reaction time differences between cue conditions, collapsing across groups, revealed that reaction times were slower to the neutral cue than the other conditions, p’s<.01, but there were no reaction time differences between the small, medium, and large reward cues.

A Group (ANH, Control) × Cue (neutral, small reward, medium reward, large reward) ANOVA on valence ratings revealed no main effect of Group or Group x Cue interaction, p’s > 0.07, but a main effect of Cue, F(3,105)=25.4, p<0.0001. Follow-up t-tests examining valence rating differences between reward cues, collapsing across groups, revealed that valence ratings differed between all four cues, p’s<.036.

A Group (ANH, Control) × Outcome (no reward, small reward, medium reward, large reward) ANOVA on valence ratings revealed no main effect of Group, p > 0.09, but a Group × Outcome interaction, F(3,105)=6.37, p<.001, and a main effect of Outcome, F(3,105)=127.8, p<0.0001. Follow-up t-tests examining valence rating differences between outcomes, collapsing across groups, revealed that valence ratings differed between all four outcomes, p’s<.0001. The Figure below illustrates task reaction times to cues and valence ratings elicited by cues and rewards.

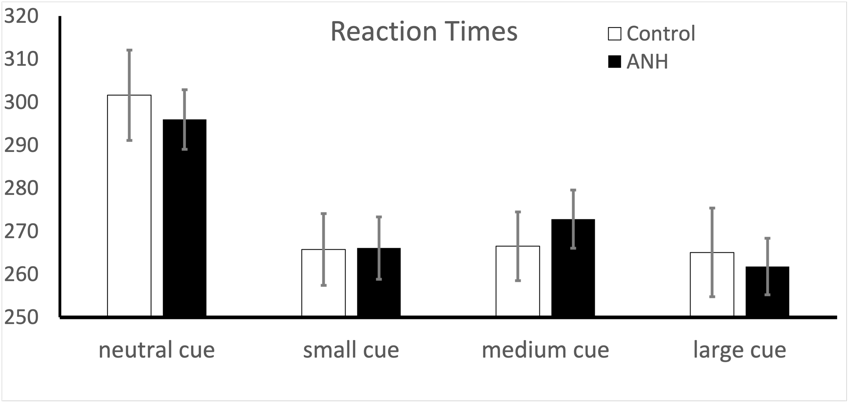

Valence Ratings of Cues	Valence Ratings of Outcomes

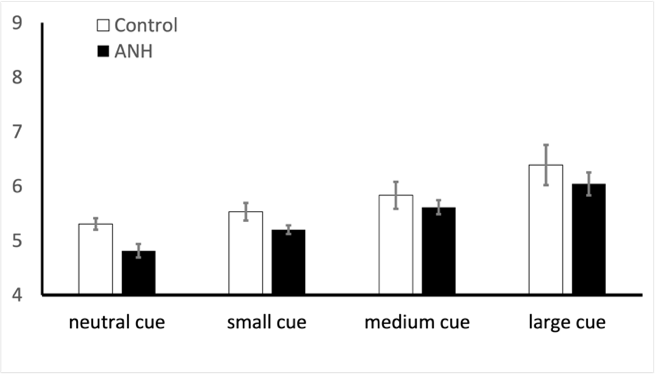

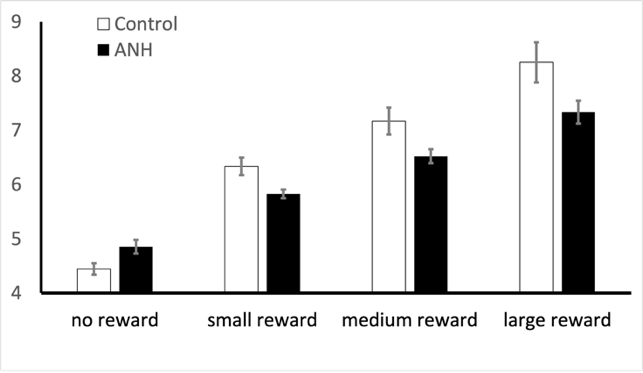

Task reaction times to cues and valence ratings elicited by cues and rewards. Valence ratings were made using a 9-point Likert scale with anchors of “very negative” (1) and “very positive” (9) at the ends and “neutral” (5) in the center. Note: the ‘neutral cue’ refers to the cue that predicted the no reward outcome, whereas the ‘small’ ‘medium’, and ‘large’ cues predicted the fifty cent, one dollar, and five-dollar outcomes, respectively. Conditions differed from each other at *: p<.05; **: p<.01; ***: p<.005. Error bars represent standard errors of the mean.

B. Absolute Difference in BP_ND_ during the MID Task (Reward – Neutral Conditions) by Groups (PET)

We first estimated BPND values by task condition (i.e., reward and neutral) only in the ANH Group (n=25). No regions showed a significant change in BPND from the neutral to reward conditions of the MID at a threshold of z > 2.58. Next, we estimated BPND values by task condition in the CON Group (n=12). Two regions in the left caudate and putamen showed a significant change in BPND from the neutral to reward conditions of the MID at a threshold of z > 2.58. A cluster of lesser significance, z < 1.5, was observed in the right putamen.

C. Group Differences in BP_ND_ during the MID Task (Reward – Neutral Conditions)

An ad-hoc binomial test indicated that the proportion of ANH participants showing an increase in BP_ND_ from Neutral to Reward conditions of the MID was greater than chance (i.e., different from 50%) in the left putamen (*p* = .032), right putamen and pallidum (*p* = .014), and left nucleus accumbens and putamen (*p* = .032) clusters. This proportion was not greater than chance in the left caudate cluster (*p* = .133).

D. Correlations between Striatal Dopamine and Functional Activation

We explored the correlations between striatal DA release and fMRI activation within the four clusters reported in the main text. These were located within the left putamen, right putamen and pallidum, left caudate, and left nucleus accumbens. We extracted the fMRI signal from these PET-derived clusters during the reward anticipation phase of the MID reward task. We found no significant correlations between fMRI activation and PET-derived *PET-derived ΔBPND values.*

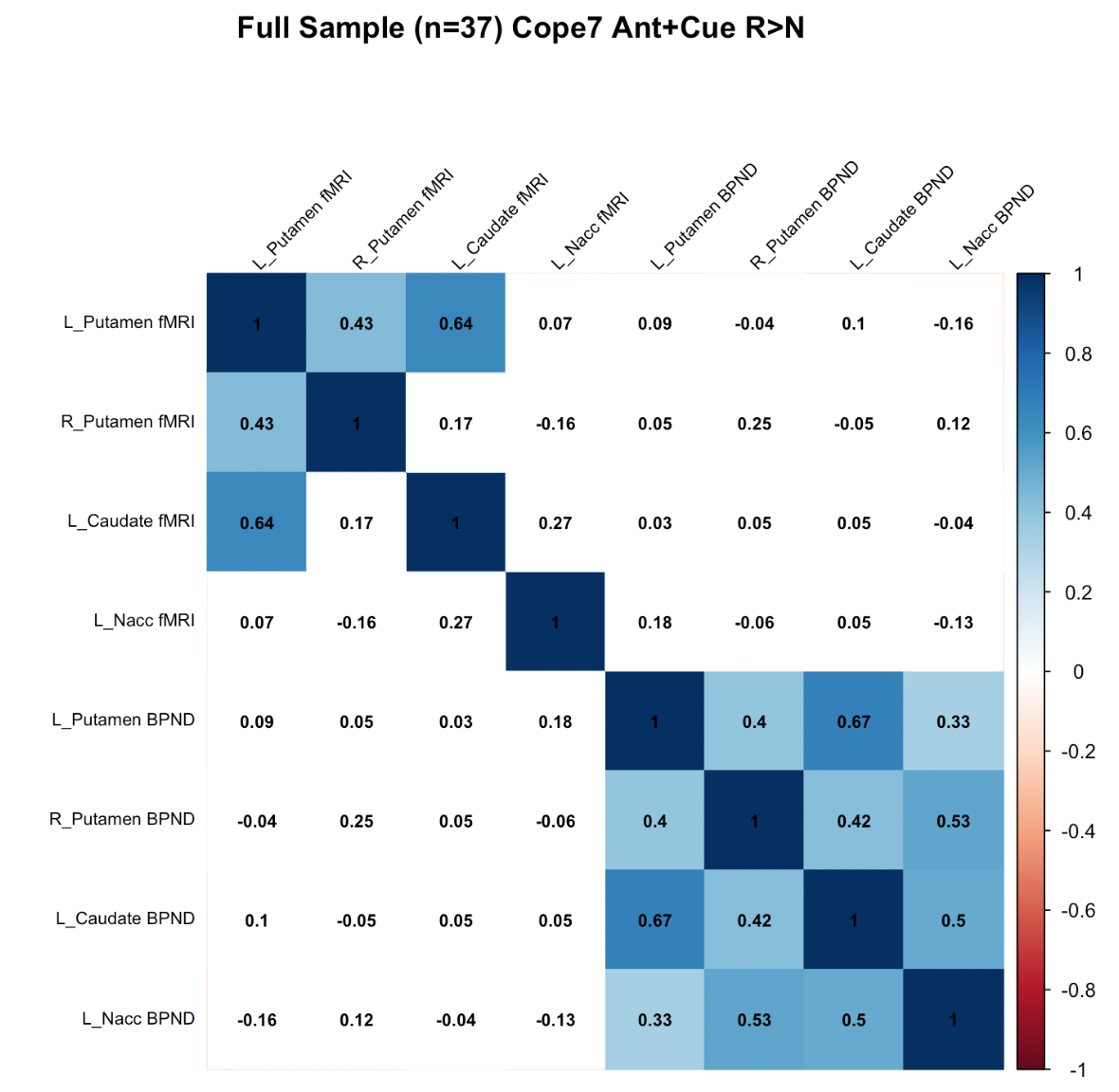

E. Striatal Dopaminergic Functioning (PET) in Structurally-defined ROIs

Structurally-defined left and right putamen ROIs derived from the AAL3 atlas demonstrated between-group differences in BP_ND_ values for the contrast of ANH - CON; Reward - Neutral. Mirroring the results reported in the main text, on average, ANH participants showed higher [11C]raclopride BP_ND_ during the reward condition of the MID relative to the neutral condition for both putamen ROIs (t’s = 0.007, p’s < .05). We also explored differences in baseline raclopride binding potential during the neutral condition. Using an ROI approach, we found no significant baseline differences between groups in binding potential in the left caudate, right caudate, and left putamen. We did find that baseline binding potential in the right putamen is significantly lower in anhedonic participants than in control participants (p<.02). Baseline binding potential also differed between groups in the left and right nucleus accumbens; however, results were in opposite directions in left and right ROIs. These NAc atlas regions are very small and may provide unreliable estimates of group differences. Our findings are mixed, and results that show no differences in baseline binding potential between groups is not consistent with a recent PET study in a clinically depressed sample (14); thus, representing a potential difference between depressed and anhedonic populations.

F. fMRI Activation

First, a whole-brain general linear model approach was used to examine whole-brain BOLD fMRI responses during reward anticipation and reward outcomes on the MID task. However, cluster-corrected results yielded no significant clusters that differentiated groups at a threshold of z > 2.58 during reward anticipation or reward outcome phases. In the Control group alone, the task reliably activated the striatum during reward anticipation. We next examined BOLD percent-signal change in mesocorticolimbic network regions-of-interest (ROIs) during reward anticipation and reward outcomes on the MID task. Across cue phases of the MID task, contrasting Reward - Neutral cues, the ANH group showed increased activation relative to the CON group in the right caudate at an uncorrected threshold, but not a corrected threshold (t(26.8) = 2.58, p = .016, pFDR = .144). Additionally, across cue phases and fixation phases (i.e., a broader anticipation window than cue phase alone, see *Figure 2* in Main Text) of the MID task, contrasting Reward - Neutral anticipation trials, the ANH group showed increased activation relative to the CON group in the right caudate at un uncorrected threshold but not a corrected threshold (t(21.0) = 2.26, p = .035, pFDR = .315). There were no group differences in activation during reward outcomes.

G. Anhedonia and Phasic DA Release in Structurally-defined Striatal ROIs (PET)

We examined the relations between dopaminergic functioning within structurally-defined striatal ROIs and anhedonia severity within the ANH group. Reduced phasic DA release to rewards in the right nucleus accumbens significantly predicted BDI-II anhedonia subscale scores (b_STD_ = .47, SE = 0.18, t = 2.58, p = .017), controlling for age and sex (Figure 4). Interestingly, there were also inverse relations between BP_ND_ values in other structurally-defined striatal ROIs and SHAPS scores: increased phasic DA release to rewards in the left caudate (bSTD = -.40, SE = 0.19, t = -2.10, p = .047) and right putamen (bSTD = -.42, SE = 0.19, t = -2.22, p = .036) significantly predicted SHAPS scores (Figure 4).

H. Anhedonia and Mesocorticolimbic Activation during Reward Anticipation (fMRI)

We examined associations between BOLD activation in the right caudate during reward anticipation (Reward - Neutral) during the MID task and anhedonia severity scores within the ANH group. Neither SHAPS nor BDI-II anhedonia subscale scores were significantly associated with BOLD activation in the right caudate during reward anticipation (p > .05).

I. Chronic Stress and Phasic DA Release to Rewards in Structurally-defined Striatal ROIs (PET)

Analyses with the PSS yielded one significant association between chronic stress and phasic DA release to rewards in striatal ROIs. Increased phasic DA release to rewards in the structurally-defined right putamen (bSTD = -.40, SE = 0.19, t = -2.10, p = .047) significantly predicted PSS scores, wherein higher PSS scores were associated with lower BP_ND_ values (i.e., increased DA release to rewards), which was the opposite of what we predicted. Exploratory analyses with our secondary measure of chronic stress, the PCL-5, yielded no significant associations between chronic stress and phasic DA release to rewards in structurally-defined striatal ROIs.

J. Chronic Stress and Mesocorticolimbic Activation during Reward Anticipation (fMRI)

Analyses with the PSS and PCL-5 yielded no significant associations between chronic stress and mesocorticolimbic activation during reward anticipation.
